# Breaking the Status Quo in Heart Failure: Leveraging Remote Patient Monitoring to Effectively Put the Heart Failure Guidelines to Practice

**DOI:** 10.1101/2023.12.11.23297939

**Authors:** David I. Feldman, Marcus L Campbell, Theodore Feldman, Randall Curnow, Marat Fudim

## Abstract

**Background:** Despite guideline directed medical therapy (GDMT) being recognized to improve outcomes in patients with heart failure with reduced ejection fraction (HFrEF), optimization has been limited resulting in worse outcomes and exorbitant costs. Although remote patient monitoring (RPM) has proven to help improve patient care, implementation of RPM at scale leading to healthcare cost savings has not been demonstrated.

**Methods:** Patients from 11 states were enrolled from August 2021 to April 2023 in a virtual heart failure (HF) program offered by Cadence. Eligible patients were Medicare beneficiaries with a history of an ejection fraction (EF) <40%. A clinical team monitored patient daily vitals measured on a cellular enabled blood pressure (BP) cuff, heart rate monitor and weight scale. Clinical visits using technology enabled clinical protocols were also conducted on a regular basis to facilitate guideline directed clinical interventions including symptom, vital and medication optimization. Cost analysis used 5 years of de-identified healthcare claims data from an Accountable Care Organization (ACO) and calculated average monthly healthcare costs using the 4-month period of January-April for each year. We then used a Differences-in-Differences analysis to estimate the effect of Cadence on average monthly healthcare costs compared to ACO patients who were ordered for Cadence but did not enroll.

**Results:** Total of 367 patients (mean [SD]: age 74 [11] years; EF 45 [2] %; systolic BP (SBP) 131 [19] mmHg; n [%]: 122 women [33%]; 260 white (71%)) were followed for a median of 294 days. There was a significant decrease in patients’ BP (SBP -6.9, Diastolic BP -4.9 mmHg; p<0.001) and weight (−2.1 lbs; p=0.010) but not heart rate. Patients experienced significant increases in the use of sodium glucose co-transporter 2 inhibitors (92 [26%] to 165 [45%]; p<0.001) and mineralocorticoid receptor antagonists (120 [33%] to 144 [39%]; p=0.002) but not β-blockers or renin-angiotensin system antagonists. The percentage of patients on ≥50% target dose significantly increased for all pillars of GDMT. There was also a significant increase in the percentage of patients on all 4 pillars of GDMT at follow (84 [23%] vs. 26 [7%]; p<0.001). A total of 70 enrolled and 42 ordered but not enrolled HF patients were included in the ACO analysis. Compared to ordered but not enrolled patients, enrollment in Cadence resulted in a 52% (-$1,076.64 per HF patient per month) cost reduction, with the most significant reductions related to hospitalizations and hospital associated spending.

**Conclusions:** We present the first evidence to support the use of a remote patient intervention program that leverages RPM and technology supported clinical interventions to not only improve the use and dose of GDMT for HFrEF patients but also reduce total and hospital associated costs.

---

Guideline directed medical therapy (GDMT) effectively reduces morbidity and mortality for patients with heart failure (HF) [1]. Unfortunately, despite clear recommendations for initiating and titrating GDMT [2], optimization of GDMT in HF patients nationwide is staggeringly low [3]. Reasons such as clinical inertia, inaccessibility of clinicians, and patient level cost and medical constraints are often to blame [4]. Remote patient monitoring (RPM) has helped address these challenges and resulted in improved care for HF patients [5]. Further adoption of comprehensive RPM and centralized medication titration by health systems is required to close care gaps, improve outcomes, and reduce cost. Outsourcing the GDMT titration efforts to a centralized RPM program could be one of the most impactful solutions.

We studied the impact of a healthcare technology company (Cadence), which leverages RPM and delivers clinical care for patients with chronic diseases alongside healthcare institutions. Medicare beneficiaries with an ICD-10 code for heart failure with reduced ejection fraction (HFrEF) and who were enrolled from August 2021 to April 2023 were included in this analysis. A remote multidisciplinary clinical team monitored patient’s daily vitals measured on a cellular enabled blood pressure cuff, heart rate monitor and weight scale. Virtual clinical visits using standardized clinical protocols were also conducted on a regular basis to facilitate guideline directed clinical interventions including symptom, vital and medication optimization. The cost analysis used 5 years of de-identified healthcare claims data in HF patients from a Cadence partner Accountable Care Organization (ACO) and calculated average monthly healthcare costs using the 4-month period of January-April for each year. Using a Differences-in-Differences analysis, we estimated the effect of enrolling in Cadence on average monthly healthcare costs compared to ACO patients who were ordered for Cadence but did not enroll.

A total of 367 patients (mean [SD]: age 74 [11] years; EF 45 [2] %; systolic blood pressure 131 [19] mmHg; n [%]: 122 women [33%]; 260 white [71%]) were followed for a median of 294 days. There was a significant decrease in patients’ blood pressure (systolic blood pressure -6.9, diastolic blood pressure - 4.9; p<0.001) and weight (−2 lbs; p=0.010) but not heart rate. Patients experienced significant increases in the use of sodium-glucose cotransporter-2 inhibitors (92 [26%] to 165 [45%]; p<0.001) and mineralocorticoid receptor antagonists (120 [33%] to 144 [39%]; p=0.002) but not beta-blockers or renin-angiotensin system antagonists. The percentage of patients on ≥50% target dose significantly increased for all pillars of GDMT **(Figure 1A)**. There was also a significant increase in the percentage of patients on all 4 pillars of GDMT at follow up (84 [23%] vs. 26 [7%]; p<0.001).

**Figure 1:**
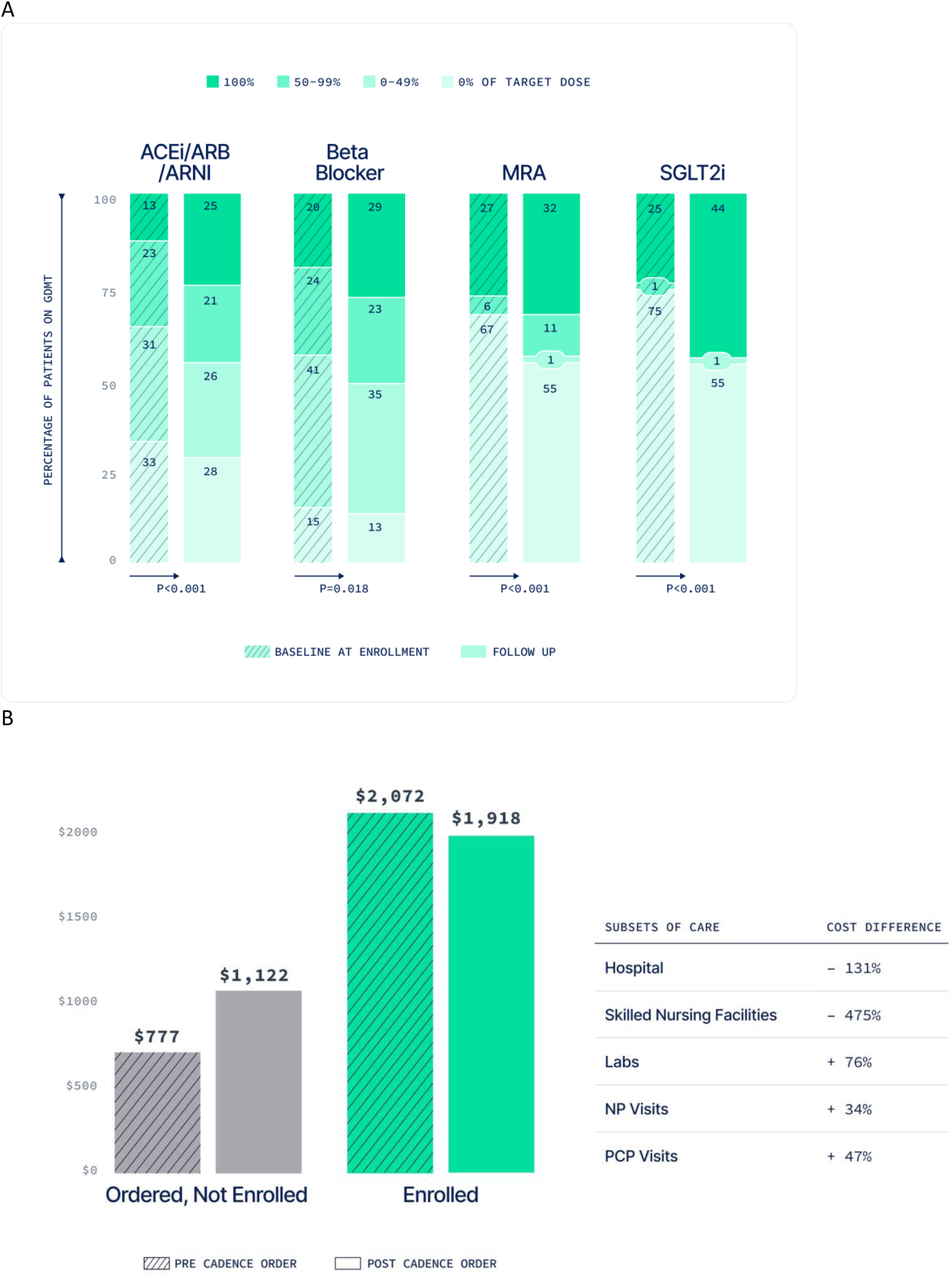
Panel A – Use and dose of GDMT at follow-up vs. baseline; Panel B – Effect of Cadence on Healthcare cost in enrolled vs. non-enrolled patients. Abbreviations: ACEi – angiotensin converting enzyme inhibitor; ARB – angiotensin receptor blocker; ARNI – angiotensin receptor-neprilysin inhibitor; MRA – mineralocorticoid receptor antagonists; SGLT2i – sodium-glucose cotransporter-2 inhibitor; GDMT – guideline directed medical therapy; NP – nurse practitioner; PCP – primary care physician.

The ACO cost analysis included a total of 70 HF patients enrolled in Cadence and 42 HF patients ordered for Cadence not enrolled. Compared to ordered but not enrolled HF patients, enrollment in Cadence resulted in a 52% reduction in total cost of care (-$1,076.64 per HF patient per month) despite an increase in cost related to primary care physician and nurse practitioner visits. Most cost savings were secondary to decreases in hospital and post hospital discharge (ie. skilled nursing facility) related spending **(Figure 1B)**.

This data highlights the benefit of a comprehensive RPM solution on improving clinical outcomes for patients with HF. In addition, it is the first data to support the effect of an end-to-end RPM solution on reducing total and hospital associated cost. Advancements in the treatment of HF have far surpassed our ability to implement these lifesaving therapies effectively. A preferred alternative to today’s homegrown GDMT clinics might include collaboration with an outside, independent RPM and virtual care program to support GDMT titrations at primary care and cardiology clinics, particularly at under resourced institutions and in communities serving patients at risk for and affected by social determinants of health. We must break the status quo of sub optimally treating patients with HF at the expense of poor clinical outcomes and exorbitant cost, and instead embrace solutions like comprehensive RPM programs with centralized medication titration that will help fill the gaps in care for patients with HF.

## Data Availability

All data produced in the present study are available upon reasonable request to the authors.

## Notes

### Competing Interest Statement

DIF and MF are advisors at Cadence; MLC, TF and RC are employees at Cadence

### Funding Statement

This study did not receive any funding

### Author Declarations

IRB of Duke University gave ethical approval for this work. IRB # Pro00114791

